# Assessment of Ultraviolet and Infrared Radiations Transmission in Automobile Windshields and Side Windows

**DOI:** 10.1101/2024.05.27.24307977

**Authors:** Nouf Jubran AlQahtani, Ghada Naje AlEssa, Hoor Fayez AlDushaishi, Amnah Nabil Bukair, Syed Mehmood Ali

## Abstract

**Background:** Although exposure to solar radiation is beneficial for humans, too much of it can cause severe health conditions, including sunburn and skin cancer. The biological effects of solar radiation vary enormously with wavelength and exposure time. Ultraviolet and infrared radiations are the two main invisible components of solar radiation, causing skin damage. People are becoming more aware of the significance of sun protection, though little attention is directed to the exposure of the skin to UV and IR radiations through car windows. According to a survey of 1293 participants, mainly from Saudi Arabia, the fact that UV radiation can penetrate through car windows is known by the majority, i.e., 82%. However, the capability of IR radiation to penetrate vehicle windows is unknown to most people. Even though car windows reduce the transmission of ultraviolet and infrared rays, drivers are not isolated from them completely. To the best of our knowledge, this is the first study that measures solar exposure in cars in the middle east region, which is famous for its hot and arid (dry) climate with temperatures reaching over 52°C. Specifically, this study aimed to determine the driver exposure to UV and IR radiations in Dammam, Saudi Arabia, and emphasize the need to take the necessary measures to avoid exposure to these rays.

**Method:** An experiment using a PMA2100 datalogger radiometer was conducted inside and outside twenty vehicles. The radiometer measured the transmitted radiation through the front and the side windows. Then, the UV and IR measurements were analyzed and evaluated inside all vehicles.

**Results:** The average transmission percentage of ultraviolet through the car’s side window was 10.47%, while the front window only transmitted 4.06%. In comparison, the average transmission percentage of infrared through the side window was 30.02% and 39.30% through the front windows.

**Conclusion:** These results highlight the importance of protection against UV and IR radiations since the drivers are exposed to significant UV and IR radiations. Furthermore, the protection methods must be increased in areas with a hot climate, such as the middle east region.

## 1 INTRODUCTION

Sunlight is a double-edged sword since life cannot be sustained without it. However, chronic exposure to sunlight is linked to the prevalence of irreversible skin damage, skin cancer, and eye disorders. Sunlight consists of three main rays, which are Ultraviolet (UV; 280–400 nm), Infrared (IR; 760 nm–1 mm), and visible light (VL; 400–760 nm; not shown in this work).^1^ These rays can alter the physiological functions of the body, leading to serious health implications. Accordingly, uncontrolled exposure to UV and IR radiations is one of the main reasons that cause the development of some disorders. For example, exposure to UV radiation is the leading cause of sunburn, skin cancer, and skin aging.^2^ On the other hand, IR radiation has a significant role in skin aging and Erythema ab igne.^3^ According to epidemiological studies, IR radiation causes skin carcinogenesis.^4^ Furthermore, IR radiation can raise the skin temperature to 43°C, then convert it into heat energy leading to severe effects on the skin tissues.^5^ Although these rays are invisible, they are destructive, especially for human skin. Such radiations interact with the skin through three mechanisms: scattering, absorption, and reflection based on skin layers, wavelength, and energy of radiation. In general, there is an inverse relationship between energy and wavelength. For instance, the shorter wavelength has lower energy than the longer wavelength; thus, limited penetration level compared to the longer wavelength. The deeper the penetration, the greater the risk. Among solar radiation components, UV radiation has the most severe effects on the human body due to its shorter wavelengths and higher energy.^6^ In contrast, IR radiation has a longer wavelength and lesser energy leading to deep penetration to the dermis layer.^1^

Many studies demonstrated the negative impacts of uncontrolled exposure to solar radiation on human health. ^7–11^ Hence, it is crucial to monitor the amount of UV and IR radiation that affect human health negatively. Two measurements were globally used to indicate the risk of UV and IR, which are UV index (UVI) and IR irradiance, respectively. UVI describes the UV level, where each range of UVI is associated with a specific level of risk and required protection. UVI is unitless and ranked from 0 (no UV radiation) to +11 (extremely dangerous radiation). UVI of three or above is sufficient to cause health implications, such as skin damage and skin cancer. ^12^ Likewise, irradiance measures solar power, which is the rate of solar energy within a surface. If the irradiance value is high, it indicates high radiation. Cumulative exposure to UV and IR is much more dangerous than a single exposure.

Even though UV and IR exposure hazards are well known, humans cannot avoid them entirely. As a result, there has been a huge trend to design protective devices and spread awareness of protection methods against dangerous exposure to solar radiation. Some examples of protection techniques against solar exposure include the application of sun protection factors (SPF) and sunglasses. Yet, these techniques offer limited protection. Thus, the idea of designing personalized monitoring devices has emerged as a potential solution. For example, Amini et al. have developed a UV monitoring embedded device. This device is used with specific software that displays the personal history of UV exposure with the dose received during the day/week/month.^13^ Moreover, there is a newly designed wearable, thin, and stretchable UV sensor used for UV and sunscreen protection quantification by Shi et al.^14^ This sensor is an ultra-thin UV patch that can be used during different real-life activities. Also, it can be worn continuously for five days because of its structure and elastic properties. However, most of the designed devices focused on UV exposure only while underestimating the negative effects of IR exposure. Besides, they cannot measure exposure inside vehicles for drivers’ protection.

Some people may be able to avoid sunlight exposure during peak times, but this is not possible for outdoor workers, such as drivers. Consequently, the vehicles’ windows should protect against UV and IR exposure by some kind of shielding. Mainly, the vehicle’s windows are made of two kinds of glass, which are laminated and tempered glass, as shown in Figure 1. Laminated glass is made of more than one glass layer with a polyvinyl butyral (PVB) layer. PVB is a synthetic non-degradable polymer used in windshields since it decreases UV radiation transmission.^15^ The tempered glass is designed to protect the drivers in accident situations since its broken pieces are too small, so it is usually used for the side and back windows of vehicles. This glass transmits more UV radiation since it lacks a PVB layer.^16^ Many studies assure that UV radiation can penetrate vehicle windows, indicating that drivers are exposed to harmful radiation while driving. ^6, 17–20^ However, these studies did not consider IR exposure inside vehicles, even if it is dangerous as UV radiation. IR poisonous rays are underestimated, and the evidence is the absence of glass that can block the IR radiation inside vehicles. A study conducted by Brian et al reported that the blockage efficiency of side windows is 25% less than the front window.^19^ Another conducted by Hampton et al. proved that UV radiation transmits through the laminated glass, even with the existence of the PVB layer. PVB layer, films, and IR coatings could affect the amount of radiation transmitted through windows. IR coatings are layers made from metals that allow IR passage at low levels while reflecting it at high levels. However, these coatings are not used in various vehicles worldwide, and their existence increases the vehicle’s cost.^21^

**Figure 1:**
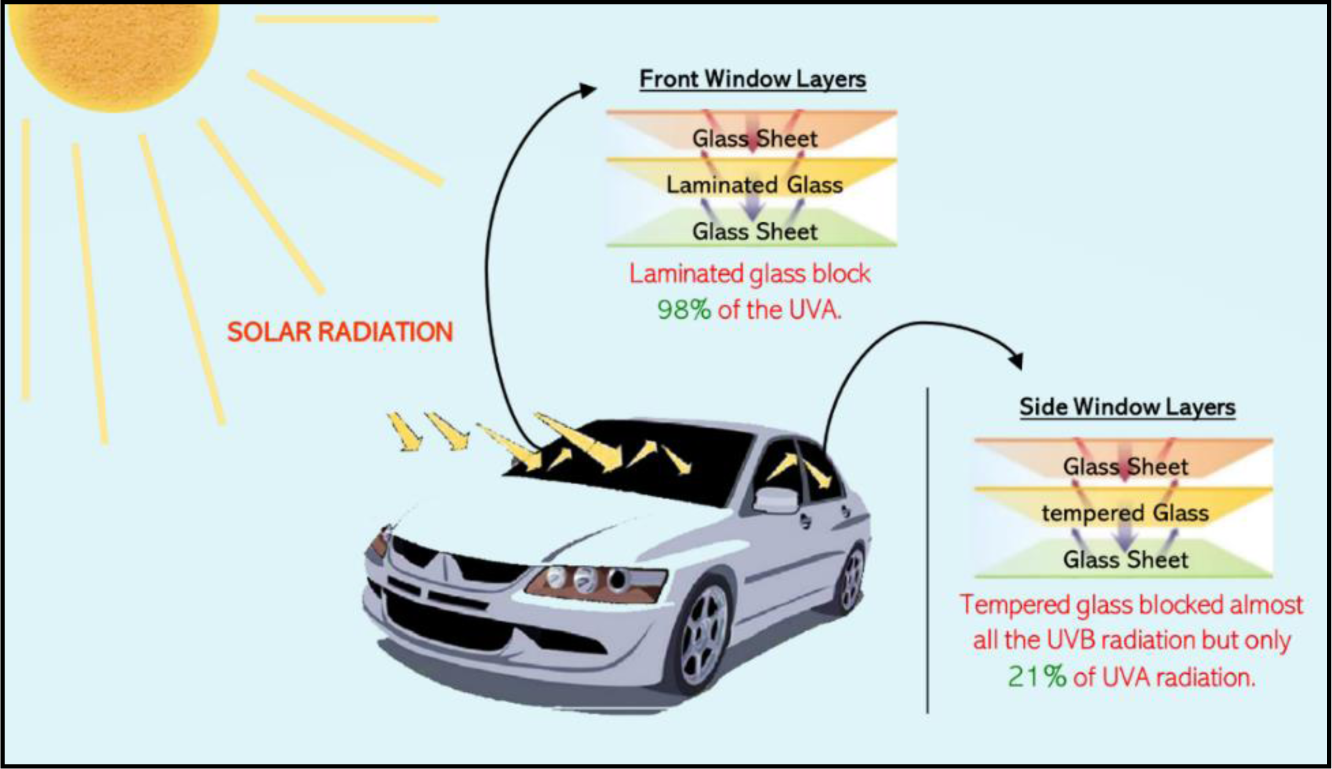
Solar radiation and car windows design^15^

Additionally, other studies proved that the driver’s part beside the window is more likely to be damaged by UV and IR radiation since it is exposed constantly to these radiations.^22–24^ Washington University researchers announced that a higher incidence of cancer was observed on the left side of the body since the study was conducted in United State (a left-driving country). In addition, they showed that skin cancer development increased in the upper left arm since it is one of the most parts exposing to the sun while driving.^22^ Furthermore, another study performed in Australia, which is a right-driving country, proved that skin cancer incidence increased on the right side.^23^

This study focuses on assessing exposure to solar radiation inside vehicles. It attempts to quantify the UV and IR protection levels in car windows. Since the drivers spend much time inside vehicles, these vehicles should protect them from adverse exposure to UV and IR radiation. It aimed to determine the extent of exposure to UV and IR radiations while driving. The data acquired from this study can be used to offer appropriate solutions to minimize solar exposure and its adverse effects. To our knowledge, this is the first study that evaluates IR exposure besides UV exposure inside vehicles. Also, it is the first study of its kind conducted in the middle east region, specifically in the Kingdom of Saudi Arabia (KSA).

## 2 METHODOLOGY

A sequential experiment was performed on twenty vehicles using Solar Light Model PMA2100 Dual-Input Data Logging Radiometer. This device can measure UV and IR radiations transmitted through car windows. One of the main steps of this work would be measuring awareness of risks associated with exposure to solar radiation. Thus, a survey was conducted, and it focused on local awareness. Afterward, the solar protection levels were assessed in twenty vehicles, where the assessment was conducted in Dammam, Saudi Arabia, from 9:00 AM. to 3:00 PM. on sunny days in June 2021. Dammam has a desert climate with an average temperature of 26.40℃ annually. Inclusion criteria in the experiment were the car windows must be original from the agency (not replaced due to accidents) and not shaded.

### 2.1 Awareness Level

Although UV and IR radiations can transmit through vehicle windows, many people do not recognize their risks and ignore precautions. Therefore, a survey was conducted to measure awareness regarding this situation. Then, data were collected to be statistically analyzed using Google sheets.

### 2.2 Exposure Measurement

Solar Light’s Model PMA2100 Dual-Input Data Logging Radiometer consists of two sensors to measure UV and IR radiations. PMA2140 sensor for detecting IR radiation, and PMA2107 sensor for detecting UV radiations. The radiometer is calibrated in terms of accuracy and precision before taking measurements. It provides readings in a unit of irradiance (mW/cm^2^). Accordingly, it is used for solar radiation measurements inside and outside the vehicle. The experiment was conducted in Dammam, Saudi Arabia, from 9:00 AM. to 3:00 PM. on sunny days in June 2021, where the temperature ranged from 44 to 46℃. The UV and IR radiations, transmitted through the front and side windows, were evaluated in 20 different vehicles, where models ranging from 2001 to 2020. One crucial factor to be considered is the position of the sensors. Therefore, the Global sensor that measures IR radiations was adjusted parallelly to the side window and the windshield. The measurements were taken from each car for sixteen minutes during a sunny and clear sky. Every minute, sensors will store one measured value. Later, data was transferred to a PC using a cable, and PMA organizer software was used to select and store a selected range of data.

### 2.3 Statistical Analysis

The data collected were statistically analyzed to identify common patterns and draw conclusions. The data obtained from the survey were analyzed using Microsoft Excel, where all data were presented graphically. Furthermore, the average of UV and IR radiation measurements are calculated for investigation purposes. Data analysis is processed descriptively. The minimum, maximum, mean, and standard deviations are determined to describe UV and IR exposure inside and outside vehicles.

## 3 RESULTS AND DISSCUSSION

### 3.1 Survey Analysis

The survey was conducted to collect data about awareness level of solar radiation exposure, where 1293 participants answered various questions. The questions involve the country of residence, awareness of the risks, and whether they avoid sunlight and apply any preventive measures. The data obtained from the survey was analyzed using graphical representations, as shown in Figure 2 and Figure 3.

**Figure 2:**
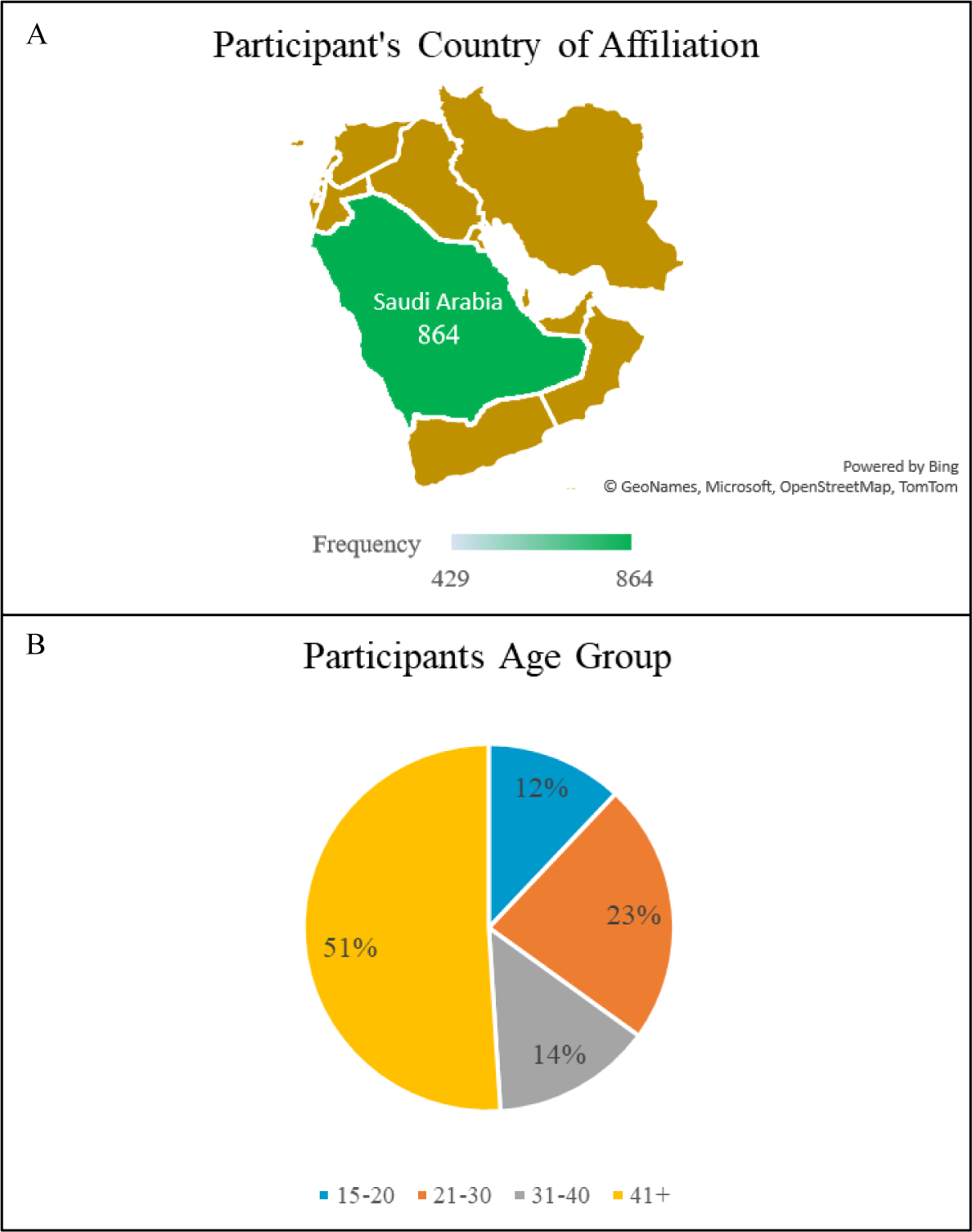
(A) Determine the geographic region of participants (B) Determine the age group of participants

**Figure 3:**
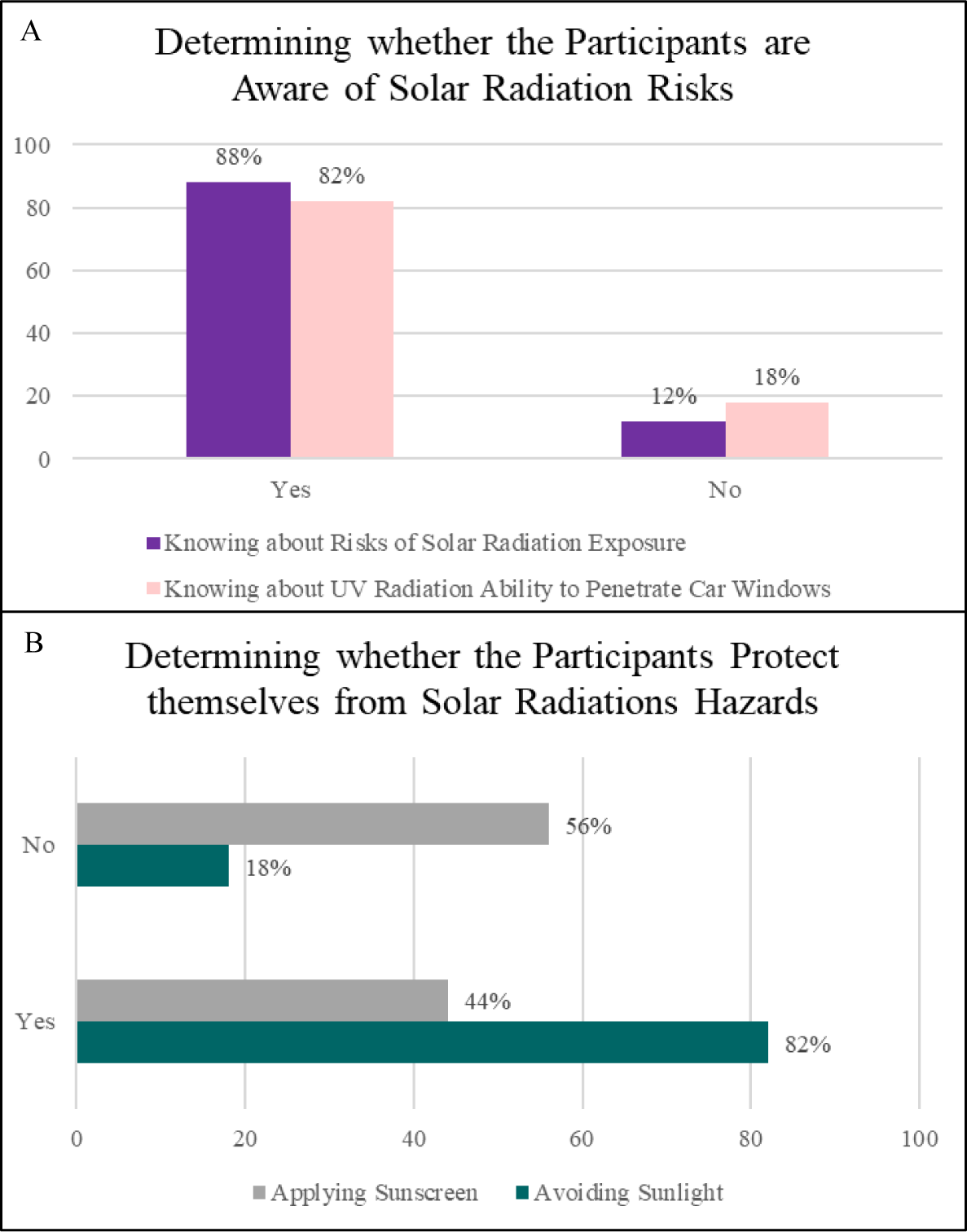
(A) Determine whether participants realize the risks of UV exposure inside and outside the vehicles (B) Determine whether participants protect themselves from solar radiation

The results showed that most participants live in Saudi Arabia with a percentage of 67% 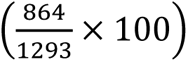, as shown in Figure 2(A). This is desirable since this study assessed exposure to UV and IR radiations inside vehicles in this country. In Figure 2(A), the frequency indicates the number of participants from inside and outside the KSA, whereas the dark green color indicates the participants from KSA. Herein, 864 participants (represented in the dark green color) were from KSA, and 429 participants (represented in the light green color) were from outside the KSA. Since the tastes and behavior of people change as they get older, the data collected from the survey included participants’ age, where 51% of the participants were over forty years old, as shown in Figure 2(B).

Even though most participants are aware of solar radiation risks, 12% of participants are unaware of risks linked to solar radiation exposure, as shown in Figure 3(A). Furthermore, the participants older than 41 showed the lowest response of unawareness of solar radiation risks. Interestingly, the age group of 31-40 showed the highest response to solar exposure risks unawareness. Only 44% of these apply sunscreen, while the majority, i.e., 82%, avoid direct exposure to sunlight, as shown in Figure 3(B). The ability of UV radiation can penetrate through vehicle windows is known by the majority, i.e., 82%. However, the capability of IR radiation to penetrate vehicle windows is unknown to most people. This is dangerous since the risk of exposure to IR radiation is no less than the risk of exposure to ultraviolet radiation.

### 3.2 Datalogger Experiment

The findings of previous studies have proven that adverse exposure to solar radiation has negative effects on human health. Yet, people expose to UV and IR radiation inside vehicles because these radiations can penetrate through car windows, increasing the chance of skin damage and skin cancer.^15^ Thus, a series of experiments were performed in Dammam, Saudi Arabia (left-hand driving country), from 9:00 AM. to 3:00 PM. during the summer. The results reported the intensity of UV and IR radiations inside and outside the vehicles. The measurements were collected using a radiometer for 16 minutes. Figure 4(A) shows that the UV radiation values increased as time passed. There was a fluctuation in IR radiation measurements between the maximum reading of 158.33 mW/cm^2^, and the minimum of 89.24 mW/cm^2^, as shown in Figure 4(B). This variation might be attributed to environmental factors, such as cloud movement. It denotes that as the clouds move, they block sunlight and absorb IR radiation, leading to variation in the IR measurements.

**Figure 4:**
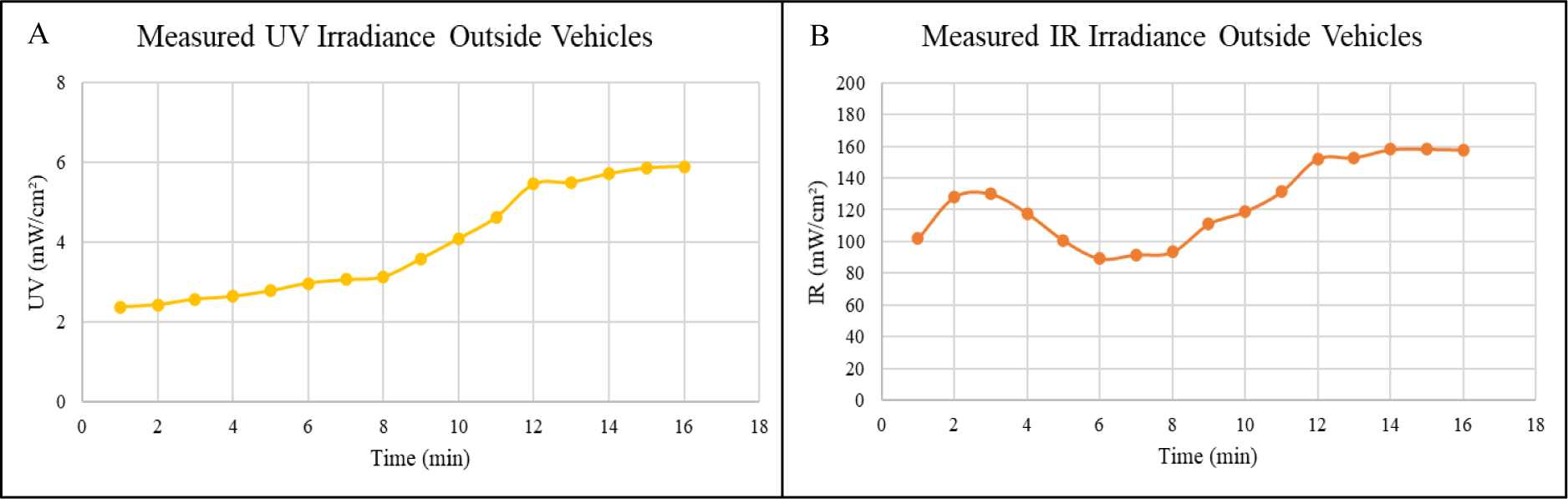
(A) UVI measurements outside the vehicles for sixteen minutes (B) IR measurements outside the vehicles for sixteen minutes

The radiometer was employed inside the vehicles to measure the UV transmitted through the front window. The Camry 2001 model transmitted the highest UV radiation through the front window, while the least amount of UV radiation was transmitted through Pajero 2015 model, where the UV radiation was 0.21 mW/cm^2^. In general, side windows transmit more UV rays than front windows because of the tempered glass used in the side windows. This type of glass allows UV rays to penetrate inside the vehicles, unlike the laminated glass used in the front windows that block most of the UV rays. Referring to Figure 5, the vehicles that transmitted the maximum UV radiations through side windows compared to others are ISUZU 2008 and Camry 2001. This may be attributed to the lack and less efficient PVB layer in the side windows of these vehicles, causing high transmission of UV rays. Besides, the ISUZU 2008 model transmitted the highest value of UV radiation through the side window, which was 1.70 mW/cm^2^, while the least amount of UV radiation was transmitted through the Denali 2013 model, where the UV radiation was 0.10 mW/cm^2^. It can be observed from the experiments that the side window can convey UV radiation more than the front window since it lacks the PVB layer. The data were statistically analyzed to draw meaningful conclusions. The analysis included the maximum, minimum, mean, and standard deviation of the UV intensities inside and outside vehicles, as illustrated in Table 1. According to data, the average exposure to UV radiation outside was approximately seven times more than inside. The measurements of the UV radiation transmitted through side and front windows in all vehicles as shown in Figure 5.

**Figure 5:**
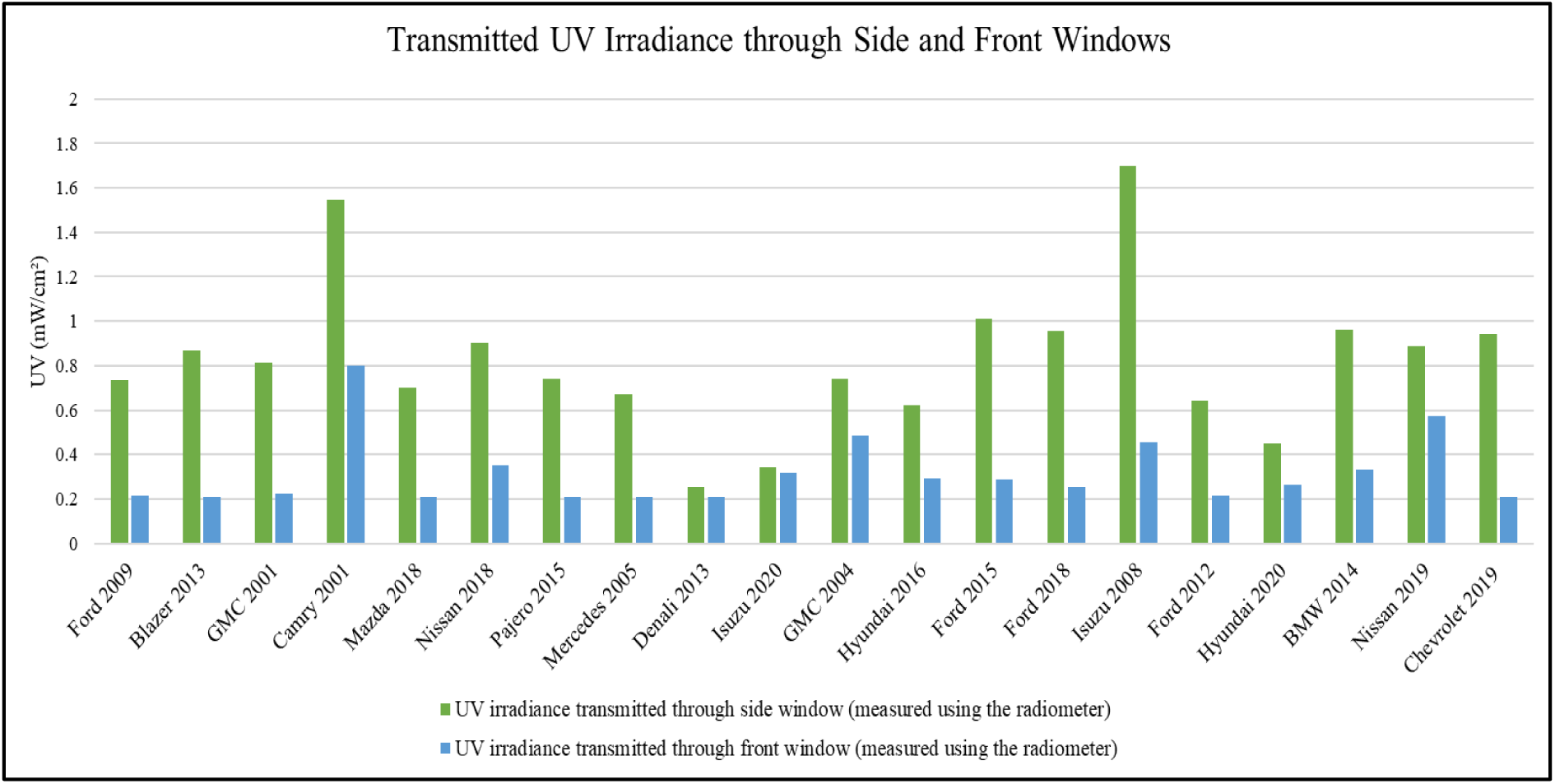
Transmitted UV irradiance through the side (represented in green color) and front (represented in blue color) windows in 20 vehicle models; the measurements were taken using the radiometer

**Table 1:** The UV intensity inside and outside the vehicle during sunny days of June 2021.

| Sampling Location | UV Intensity (mW/cm<sup>2</sup>) |  |  |  |
| --- | --- | --- | --- | --- |
|  | Maximum | Minimum | Mean | Standard Deviation |
| Outside Vehicle | 5.90 | 2.39 | 3.92 | 1.37 |
| Inside Vehicle (Front Window) | 0.80 | 0.21 | 0.32 | 0.15 |
| Inside Vehicle (Side Window) | 1.70 | 0.26 | 0.82 | 0.34 |

Likewise, IR radiation was measured in the 20 vehicles through both side and front windows. Table 2 shows the maximum, minimum, mean, and standard deviation of the IR intensities inside and outside twenty cars. The average exposure to IR radiation outside was approximately 2.3 times more than inside. The Isuzu model 2008 transmitted maximum IR radiation of 98.27 mW/cm^2^, compared to other vehicles, through the front window. On the other hand, Pajero 2015 transmitted maximum IR radiation of 84.17 mW/cm^2^ through side windows. The measurements of the IR radiation transmitted through the side and front windows in all vehicles are shown in Figure 6. In general, IR rays can penetrate side and front windows if the IR coating is not considered in these windows. As mentioned previously, IR coatings are rarely used in many vehicles worldwide as it increases the costs of the vehicles.^21^ Referring to Figure 6, some vehicles transmitted higher IR radiations through the front window than the side window due to the lack and less efficiency of IR coatings used in the front window compared to the side window. These vehicles include Isuzu 2008, Hyundai 2016, Denali 2013, GMC 2004, and Chevrolet 2019. Since there were no previous studies discussing the effect of IR exposure, we could not compare our results with previous studies. Nevertheless, our results are consistent with hypotheses and theoretical studies that prove the ability of IR to penetrate car windows, especially if IR coatings are not considered. In addition, usually, the predominant focus when manufacturing these cars is on blocking the UV rays of the sun due to their deep association with the occurrence of skin cancer. Thus, IR rays can penetrate the car windows more than UV rays.

**Figure 6:**
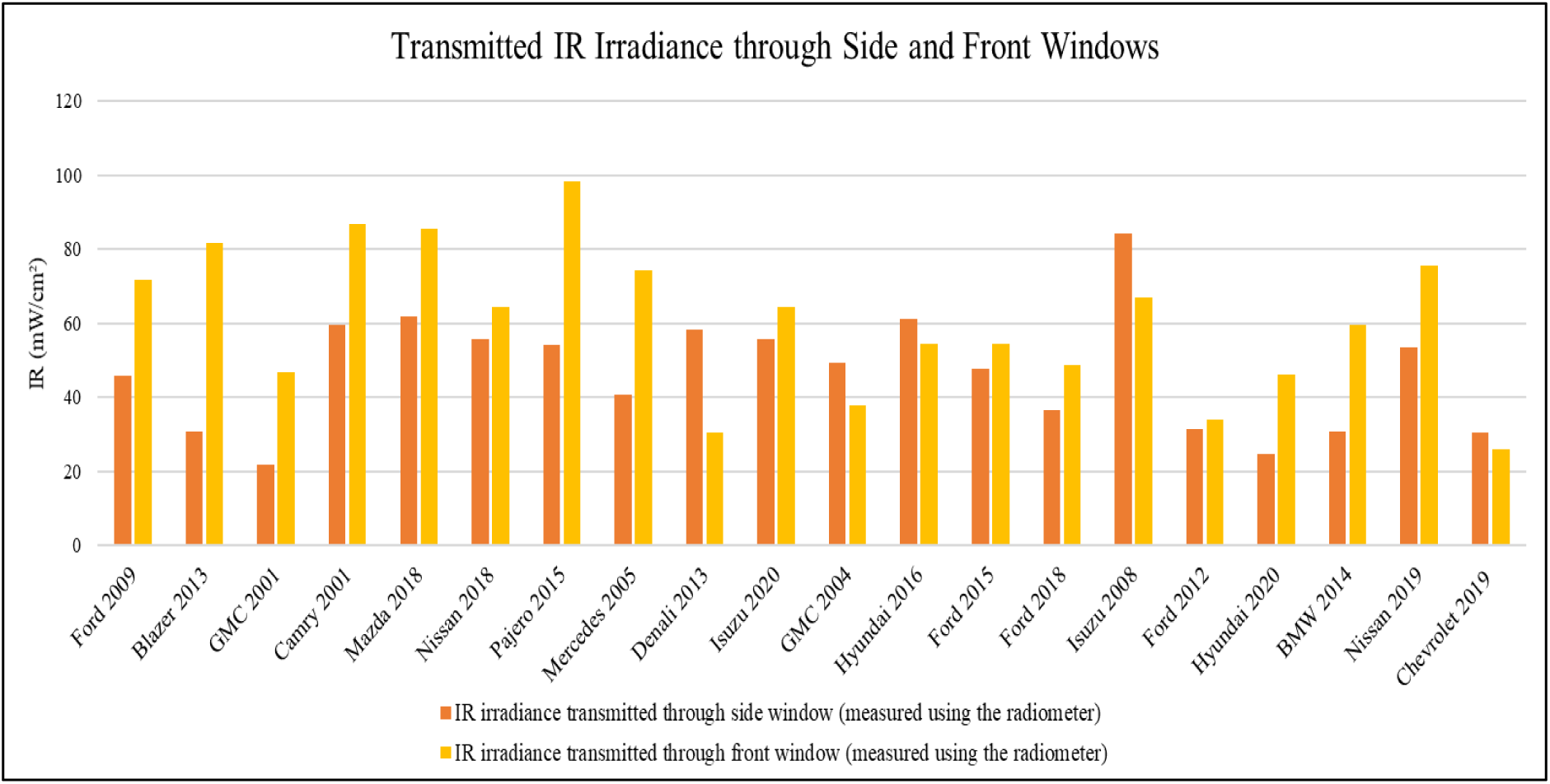
Transmitted IR irradiance through the side (represented in orange color) and front (represented in yellow color) windows in 20 vehicle models; the measurements were taken using the radiometer

**Table 2:**
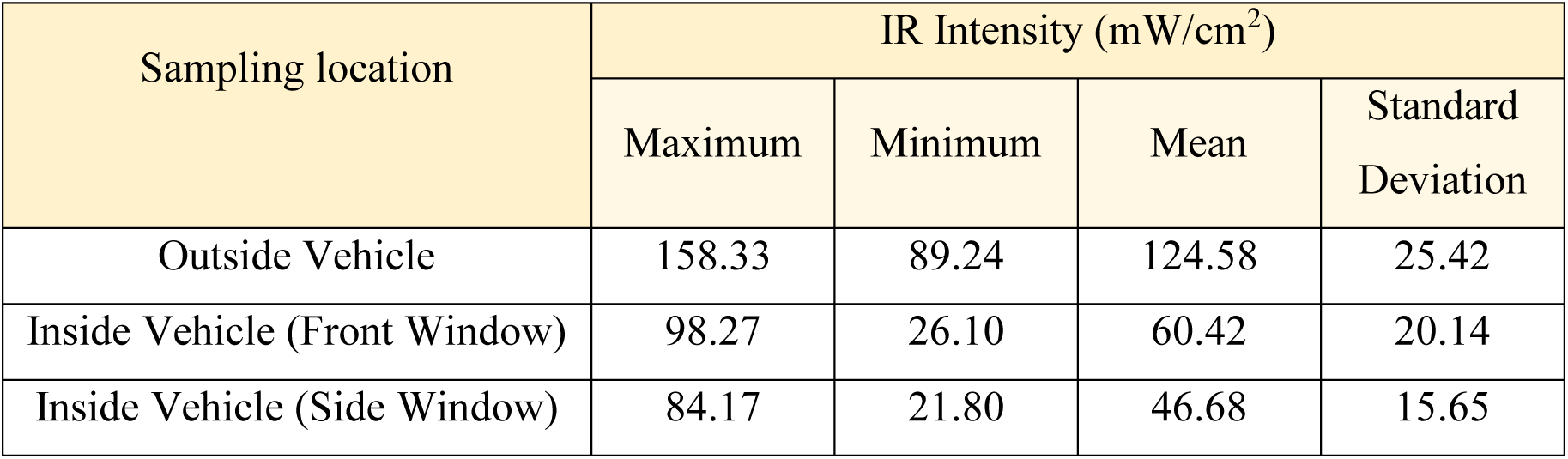
The IR intensity inside and outside the vehicle during sunny days of June 2021.

Table 3 demonstrates UV and IR measurments inside 20 vehilces besides outside measurments.

**Table 3:** The data of car experiments obtained from datalogger for 20 vehicles.

| Vehicle Model | Transmitted UV Irradiance (mW/cm<sup>2</sup>) |  |  | Transmitted IR Irradiance (mW/cm<sup>2</sup>) |  |  |
| --- | --- | --- | --- | --- | --- | --- |
|  | Outside | Front Window | Side Window | Outside | Front Window | Side Window |
| Ford 2009 | 5.90 | 0.21 | 0.74 | 157.90 | 71.87 | 45.82 |
| Blazer 2013 |  | 0.21 | 0.87 |  | 81.51 | 30.78 |
| GMC 2001 |  | 0.22 | 0.82 |  | 46.77 | 21.80 |
| Camry 2001 |  | 0.80 | 1.55 |  | 86.76 | 59.45 |
| Mazda 2018 |  | 0.21 | 0.70 |  | 85.50 | 61.72 |
| Nissan 2018 |  | 0.35 | 0.90 |  | 64.35 | 55.87 |
| Pajero 2015 |  | 0.21 | 0.74 |  | 98.27 | 54.02 |
| Mercedes 2005 |  | 0.21 | 0.67 |  | 74.22 | 40.86 |
| Denali 2013 |  | 0.21 | 0.26 |  | 30.48 | 58.22 |
| Isuzu 2020 |  | 0.32 | 0.34 |  | 64.51 | 55.66 |
| GMC 2004 |  | 0.49 | 0.74 |  | 37.69 | 49.20 |
| Hyundai 2016 |  | 0.29 | 0.62 |  | 54.59 | 61.05 |
| Ford 2015 |  | 0.29 | 1.01 |  | 54.59 | 47.74 |
| Ford 2018 |  | 0.26 | 0.95 |  | 48.84 | 36.42 |
| Isuzu 2008 |  | 0.46 | 1.70 |  | 67.02 | 84.17 |
| Ford 2012 |  | 0.21 | 0.64 |  | 33.92 | 31.37 |
| Hyundai 2020 |  | 0.26 | 0.45 |  | 46.22 | 24.86 |
| BMW 2014 |  | 0.33 | 0.96 |  | 59.70 | 30.68 |
| Nissan 2019 |  | 0.57 | 0.89 |  | 75.46 | 53.43 |
| Chevrolet 2019 |  | 0.21 | 0.94 |  | 26.10 | 30.38 |
| **Average** |  | 0.32 | 0.82 |  | 60.42 | 46.68 |

The percentages of UV and IR amounts transmitted inside vehicles are shown in Figure 7 and Figure 8, respectively. Camry model 2001 car recorded the highest percentage of UV radiation transmitted through the front, while Isuzu model 2008 vehicles had the highest UV radiation percentage through the side window. The percentage of UV transmission through the front window in the Camry model 2001 was 10.22%, and 21.75% through the side window in the Isuzu model 2008. Similarly, the percentages of IR radiation transmitted into vehicles were higher than UV radiation because it does not contain a particular layer that blocks IR; however, the side window blockage is more than the front window. Furthermore, the high percentages of IR radiation transmission through the front and side windows are 62.06% and 53.16% for the Pajero model 2015 and Isuzu model 2008, respectively. These percentages indicate the amount of rays entering the vehicles divided by the amount of rays reaching the earth, and then multiplying the result by 100. Furthermore, if the calculated percentage reaches 100%, it means that all rays reaching the earth are entering the car. This case is possible only if the windows are opened. The observed results are in agreement with prior studies that proved the ability of UV radiation to penetrate vehicles’ windows.^6, 17, 20^

**Figure 7:**
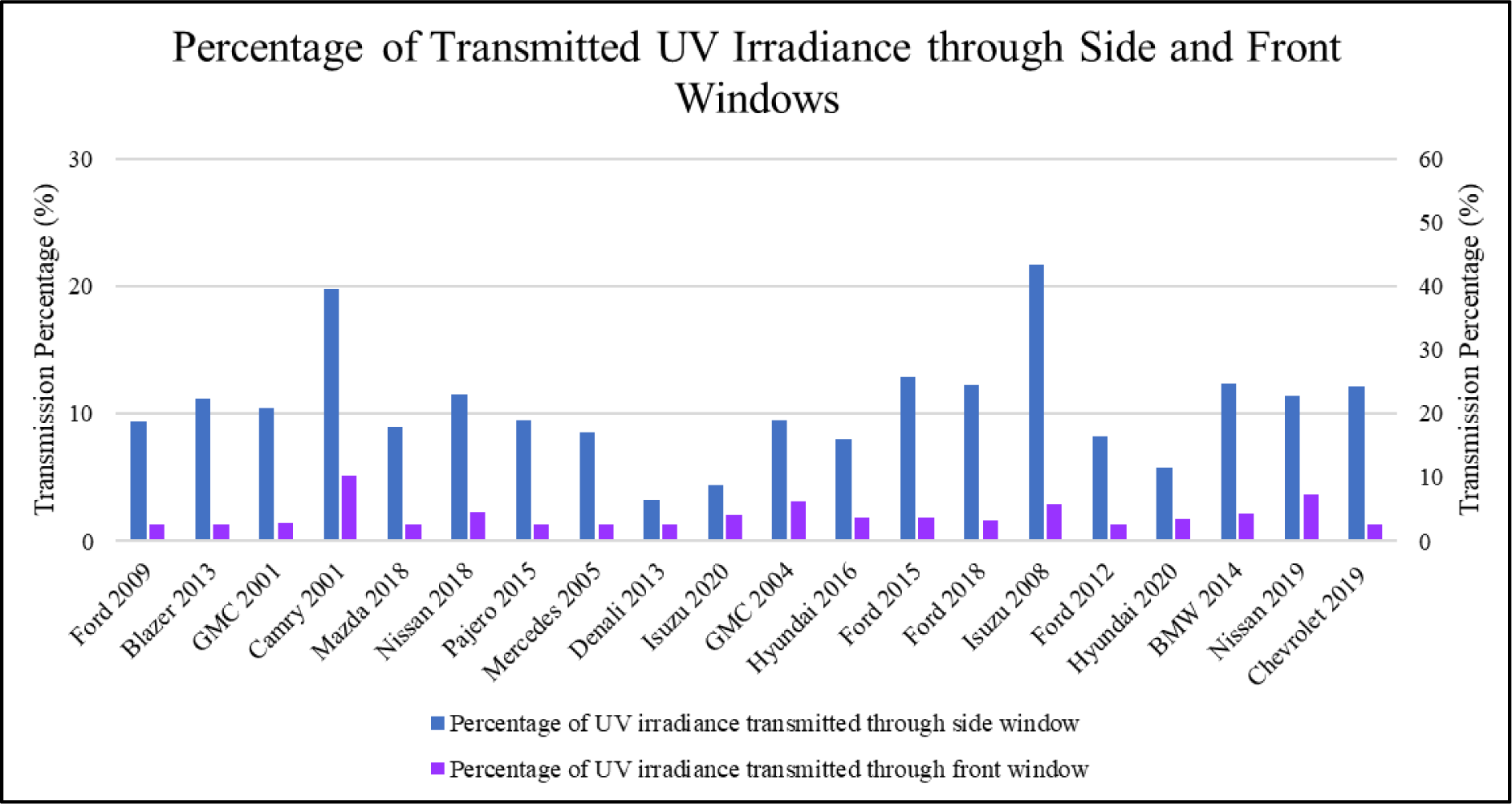
Percentages of transmitted UV irradiance through vehicle’s windows. Each car model has two columns, the blue represents transmitted UV value through the side window, while the purple represents transmitted UV value through the front window

**Figure 8:**
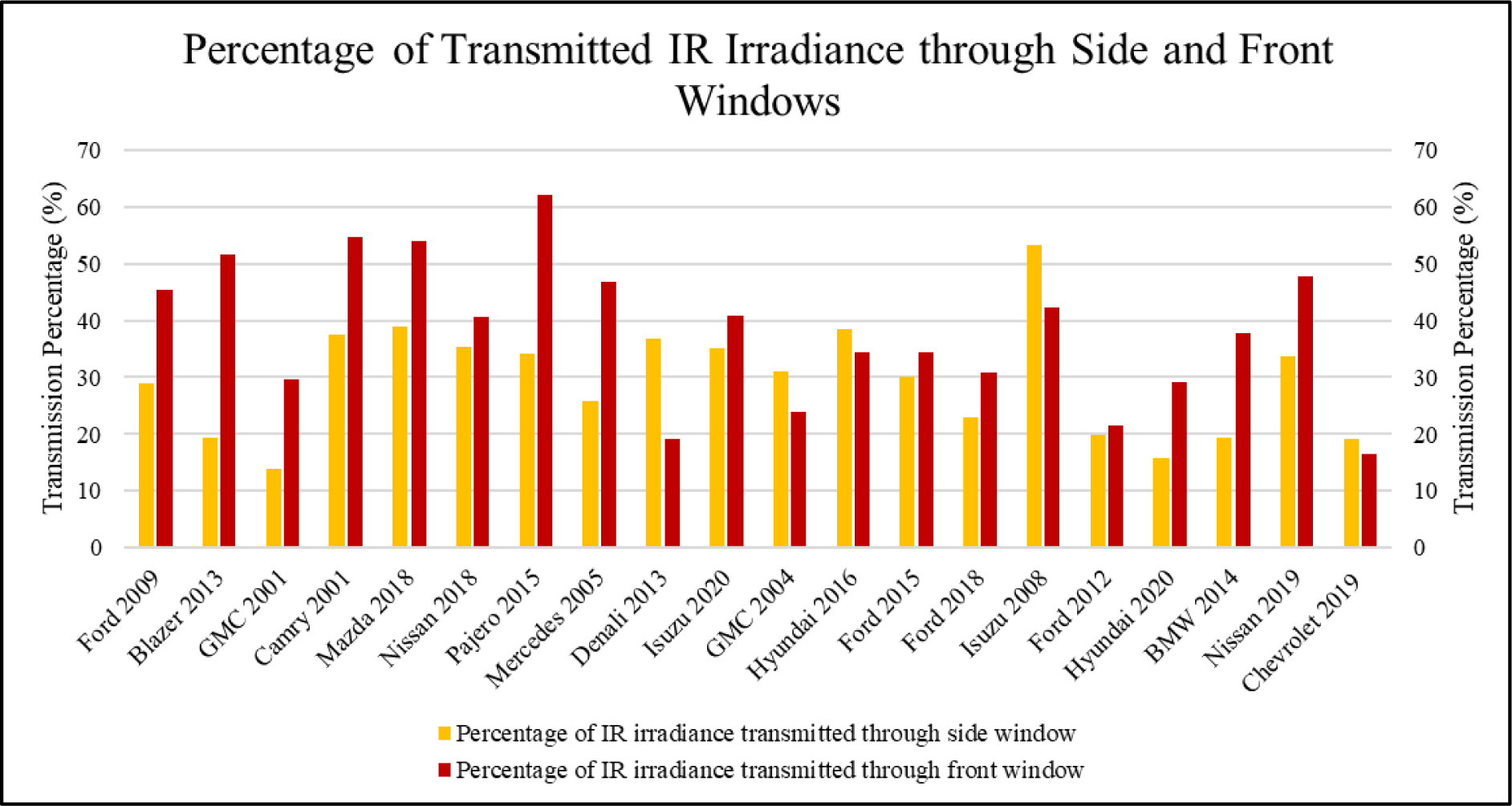
Percentages of transmitted IR irradiance through vehicle’s windows. Each car model has two columns, the yellow represents transmitted IR value through the side window, while the red represents transmitted IR value through the front window

The Isuzu model 2008 transmitted the highest amount of UV radiation in the side windows; this may occur due to the insufficient ability of the PVB layer in glass to block UV radiation. On the other hand, the average transmission percentage of UV through the 20 vehicles’ side windows was 10.47%, while the front window only transmitted 4.06%. The variation between the two windows might be attributed to different manufacturers, materials, and windows design. The front window is composed of Laminated glass made of multiple glass layers with PVB, which absorbs most of the UV. The side window is made of Tempered glass, which does not include any interlayer or any specific material or layer protected from UV radiation.^15^ Furthermore, the average transmission percentage of IR radiation through the side window was 30.02% and 39.30% through the front windows. These percentages are too high meaning that the drivers are not safe inside vehicles. Despite the fact that many people care about UV radiation too much and ignore IR radiation, IR radiation has the power to induce permanent damage to humans. Even though knowledge of the effects of IR radiation on skin aging is limited, recent research demonstrates that prolonged exposure to IR induces irreversible skin damage leading to premature skin aging.^5^ Even low-level IR absorption can cause damage, such as skin redness and sunburn. These results highlight the ability of IR radiation to penetrate through car windows which are not mentioned in prior studies. The current assessment study concentrates on UV and IR exposure risk determination inside vehicles. Besides, it shows the importance of considering the ability of IR penetration when designing car windows.

## 4 CONCLUSION

Although the importance of the sun is well known, it transmits some harmful UV and IR radiation to the earth. Continuous exposure to these radiations causes skin damage, such as sunburn, photoaging, and skin cancer. This study presents the ability of UV and IR radiations to penetrate vehicles’ windows. It is evaluated the protection level inside cars and how it differs depending on used materials and manufacturers. Besides, it highlights the demand for presenting an appropriate solution to protect humans from UV and IR radiation. The datalogger radiometer was used to analyze the UV and IR radiation in 20 cars. The results proved that UV and IR radiations inside vehicles are high enough to introduce skin damage, especially with cumulative exposure. The amount of IR radiation transmitted inside cars was much higher than the UV radiation amount. The percentages of IR radiation transmission through the front and side window reached 62.06% and 53.16% in the Pajero model 2015 and the Isuzu model 2008, respectively. While the highest percentages of UV radiation transmitted through the front window and side windows were 10.22% and 21.75% in the Camry model 2001 and Isuzu model 2008, respectively. Future work includes investigating a method that can protect people inside vehicles from harmful radiation.

## ETHICS APPROVAL AND CONSENT TO PARTICIPATE

Not applicable.

## CONSENT FOR PUBLICATION

Not applicable.

## DATA AVAILABILITY

The datasets during and/or analysed during the current study available from the corresponding author on reasonable request.

## AUTHORS’ CONTRIBUTIONS

All the authors have found the gaps in the literature, established the research hypotheses, conducted the experiment, and collected the data. All the analyses performed on this investigation were carried out by Nouf Jubran AlQahtani and Ghada Naje AlEssa. Syed Mehmood Ali and Amnah Nabil Bukair acquired the funds for the research. The methodology followed here was designed by Amnah Nabil Bukair and Hoor Fayez AlDushaishi. The original article is written by all authors, whereas Nouf Jubran AlQahtani reviewed and edited the manuscript.

## ACKNOWLEDGMENTS

We would like to thank the Deanship of Scientific Research (DSR) at Imam Abdulrahman bin Faisal University for supporting and funding this research (Project Number: 2020-033-Eng). Special thanks to our research supervisor Dr. Syed Mehmood Ali, for his support throughout this investigation. Also, this project could not have been accomplished without the support and help of our families, friends, and neighbors.

